# Identification and validation of novel candidate risk genes in endocytic vesicular trafficking associated with esophageal atresia and tracheoesophageal fistulas

**DOI:** 10.1101/2021.07.18.21260699

**Authors:** Guojie Zhong, Priyanka Ahimaz, Nicole A. Edwards, Jacob J. Hagen, Christophe Faure, Paul Kingma, William Middlesworth, Julie Khlevner, Mahmoud El Fiky, David Schindel, Elizabeth Fialkowski, Adhish Kashyap, Sophia Forlenza, Alan P. Kenny, Aaron M. Zorn, Yufeng Shen, Wendy K. Chung

## Abstract

Esophageal atresias/tracheoesophageal fistulas (EA/TEF) are rare congenital anomalies caused by aberrant development of the foregut. Previous studies indicate that rare or *de novo* genetic variants significantly contribute to EA/TEF risk, and most individuals with EA/TEF do not have pathogenic genetic variants in established risk genes. To identify novel genetic contributions to EA/TEF, we performed whole genome sequencing of 185 trios (probands and parents) with EA/TEF, including 59 isolated and 126 complex cases with additional congenital anomalies and/or neurodevelopmental disorders. There was a significant burden of protein altering *de novo* coding variants in complex cases (p=3.3e-4), especially in genes that are intolerant of loss of function variants in the population. We performed simulation analysis of pathway enrichment based on background mutation rate and identified a number of pathways related to endocytosis and intracellular trafficking that as a group have a significant burden of protein altering *de novo* variants. We assessed 18 variants for disease causality using CRISPR-Cas9 mutagenesis in *Xenopus* and confirmed 13 with tracheoesophageal phenotypes. Our results implicate disruption of endosome-mediated epithelial remodeling as a potential mechanism of foregut developmental defects. This research may have implications for the mechanisms of other rare congenital anomalies.

## Introduction

Esophageal atresia (EA) is a congenital abnormality of the esophagus, co-occurring with tracheoesophageal fistula (TEF) in 70-90% cases^1; 2^. The overall worldwide incidence of EA/TEF is 2.4 per 100,000 births^3^. Approximately 55% of individuals with EA/TEF are complex with additional congenital anomalies^3^ in the cardiovascular, musculoskeletal, urinary, gastrointestinal, or central nervous system^4^. The genetic causes of EA/TEF include chromosome anomalies or variants in genes involved in critical developmental processes that are dosage sensitive^5^. Several EA/TEF risk genes include the transcriptional regulators *SOX2*, *MYCN*, *CHD7*, *FANCB* and members of FOX transcription factor family^2; 5^.

Mouse models have demonstrated that precise regulation of the transcription factors Nkx2-1, Sox2, and Foxf1 by WNT, bone morphogenetic protein 4 (BMP4), and Hedgehog signaling pathways is required for patterning of the fetal foregut and separation of the esophagus and trachea^3; 6–9^. Moreover, EFTUD2 haploinsufficiency leads to syndromic EA, emphasizing the necessity of mRNA maturation through the spliceosome complex for normal development^10^. Recently we have shown that *de novo* variants are major contributors to EA/TEF genetic risk, especially in genes that are targets of SOX2 or EFTUD2^11^. However, it remains unclear how developmental signaling pathways, transcription factors, and RNA metabolism control the cellular behavior of tracheoesophageal morphogenesis.

Despite previous studies of the genetics in several syndromes that include EA/TEF and mouse models, the etiology in most cases of EA/TEF is still unexplained. To identify novel genetic etiologies of EA/TEF, we performed whole genome sequencing (WGS) of 185 individuals with EA/TEF, most without a family history of EA/TEF, and their biological parents. We confirmed our previous results from a smaller EA/TEF cohort, demonstrating an overall enrichment of *de novo* coding variants in complex cases. Functional enrichment analysis identified a striking convergence of putative risk genes in biological pathways related to endocytosis, membrane dynamics, and intracellular transport. We then used CRISPR-generated *Xenopus* mutant models to successfully confirm 13 of 18 candidate risk genes for EA/TEF. Together with recent reports that endosome-mediated membrane remodeling is required for tracheoesophageal morphogenesis in animal models^12^, this suggest that disruptions in endosome-trafficking may be a feature of many complex EA/TEF cases.

## Methods

### Patients Recruitment

Individuals with EA/TEF were recruited as part of the CLEAR consortium from Columbia University Irving Medical Center (CUIMC) in New York, USA, Centre Hospitalier Universitaire Sainte-Justine in Montreal, Canada, Cincinnati Children’s Hospital, in Ohio, USA, Cairo University General Hospital in Cairo, Egypt, University of Texas- Southwestern Medical Center in Texas, USA and Oregon Health and Science University in Portland, USA. Participants eligible for the study included those diagnosed with EA/TEF without an identified genetic etiology based upon medical record review. All participants provided informed consent. The overall study was approved by the Columbia University institutional review board and each affiliated site. Blood and/or saliva samples were obtained from the probands and both biological parents. A three-generation family history was taken at the time of enrollment, and clinical data were extracted from the medical records and by patient and parental interview.

We performed whole genome sequencing (WGS) on 185 probands diagnosed with EA/TEF and their parents. DNA from 75 probands was isolated from saliva samples, and DNA from the remaining 110 probands was isolated from blood samples. Patients with only EA/TEF were classified as isolated cases (59 in total) and patients with other type of congenital abnormalities or neurodevelopmental disorders were classified as complex cases (126 in total, Table S1).

### WGS analysis

We identified *de novo* coding variants using previously published procedures with heuristic filters^11; 13; 14^ augmented with in silico confirmation by DeepVariant^15^ (Table S2). We used ANNOVAR and VEP to annotate variants with population allele frequency^16; 17^ (gnomAD and ExAC), protein-coding consequences, and predicted damaging scores for missense variants. Variants were classified as LGD (likely gene disrupting, including frameshift, stop gained/lost, start lost, splice acceptor/donor and splicing damage variants (spliceAI^18^ DS score ≥ 0.8)), missense or synonymous. In frame deletions/insertions (multiple of 3 nucleotides) and other splice region variants were excluded in the following analysis. Variants in olfactory receptor genes, HLA genes or MUC gene family were filtered out of further analysis.

### Burden test

We divided patients into two categories based on their phenotypes (isolated and complex) and performed burden tests on both groups and the aggregated group. For each group, we divided *de novo* coding variants into four types: synonymous, LGD, missense, protein altering (defined as combination of LGD and missense variants). For each variant type, we calculated the expected number of variants based on a background mutation rate model^19; 20^. We used a single-sided Poisson test to test whether the number of observed *de novo* variants is significantly higher than expected. We performed the test in all genes, genes intolerant of loss of function variants (“constrained genes” based on gnomAD^16^ pLI≥0.5), and non-constrained genes. Population attributable risk (PAR) was calculated as: 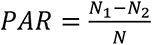, where *N*_1_, *N*_2_, *N* are the observed number of carriers of protein altering variants, expected number of carriers of protein altering variants, and the number of all cases, respectively.

### Pathway enrichment analysis of *de novo* protein altering variants in complex cases

To identify the pathways associated with *de novo* protein altering variants, we performed pathway enrichment analysis on the GO pathways and HPO terms from GSEA^21; 22^ database (version v7.2) in complex cases. We only considered the pathways with at least two protein altering variants (defined by combination of LGD and missense variants) expected by chance based on background mutation rate model^19; 20^. Based on these criteria, we selected a total of 907 pathways for downstream analysis. We performed a one-sided Poisson test of observed variants versus expectation in each pathway. Since many pathways have shared genes, we performed simulations under the null hypothesis to estimate the Family Wise Error Rate (FWER) for a given p-value. In each round, we randomly generated *de novo* LGD or missense variants based on the background mutation rate and calculated p-values for each gene. Based on simulation results, we estimated FWER by:

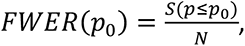

where *S*(*p* ≤ *p*0) is the total number of pathways that have p-values smaller or equal to *p*_0_ in all simulations, and *N* is the number of simulations. We used both Jaccard Index and correlation to show the overlap of two pathways. For each pair of pathways, the Jaccard Index was defined as the aggregated mutation rate of overlapping genes divided by aggregated mutation rate of all genes, and correlations were calculated as the Pearson correlation during simulation.

Network layout is generated by “Prefuse Forced Directed OpenCL Layout” algorithm in Cytoscape.

### Protein-protein interaction analysis

We tested protein interactions of *de novo* protein altering variants in complex cases using StringDB (v11.0)^23^ with default settings and default interaction sources. Edges were filtered by stringDB score≥0.4 and visualized by Cytoscape^24^. Proteins that are not connected to any other genes after interaction filtration were removed from the network. Network layout was generated by “Prefuse Forced Directed OpenCL Layout” algorithm in Cytoscape. For each gene, Degree was calculated as the sum of all stringDB scores.

### F0 *Xenopus tropicalis* CRISPR-Cas9 mutagenesis screen

All *Xenopus* experiments were performed using guidelines approved by the CCHMC Institutional Animal Care and Use Committee (IACUC 2019-0053). *Xenopus tropicalis* adult frogs were purchased from NASCO (USA) or raised in house and maintained in the CCHMC vivarium under normal housing conditions. *Xenopus* embryos were obtained by *in vitro* fertilization or natural mating as previously described^25; 26^. Germ line *sox2^-/-^* embryos (F2 generation) were obtained by mating sox2^+/-^ adults obtained from the National *Xenopus* Resource (NXR, USA; RRID:SCR_013731).

For F0 CRISPR-Cas9 indel mutagenesis, guide RNAs (gRNAs) were designed using CRISPRScan^27^ based on the Xenopus tropicalis v9.1 genome assembly on Xenbase^28^. gRNAs were designed to generate either null mutations (early in the coding sequence) or in the coding region similar to the patient mutation. *In vitro* transcribed gRNAs were synthesized using MEGAshortscript T7 Transcription Kit (ThermoFisher, USA) according to manufacturer’s instructions, or purchased as AltR-crRNA (Integrated DNA Technologies, USA). CrRNAs were annealed with AltR-tracrRNA prior to embryo injections according to manufacturer’s guidelines. Guide RNAs (500-700 pg) were complexed with recombinant Cas9 protein (1ng, PNA Biosciences) and injected into *X. tropicalis* embryos at the one- or two-cell stage. For negative controls, a gRNA designed targeting tyrosinase (*tyr*) was injected to calculate a baseline percentage of defective tracheoesophageal development in Xenopus (∼2%).

Three-day-old injected tadpoles (stage NF44) were fixed and processed for wholemount immunostaining as previously described^12^ using the following primary antibodies: mouse anti- SOX2 (Abcam, ab79351, 1:1000), goat anti-FOXF1 (R&D systems, AF4798, 1:300), and rabbit anti-NKX2-1 (SCBT, sc-13040X, 1:300). Imaging was performed using a Nikon A1 inverted LUNA confocal microscope with constant laser settings for all embryos. Image analysis was performed using NIS Elements (Nikon, USA). After image analysis each embryo was genotyped by PCR amplification of the target region followed by Sanger sequencing. Since F0 CRISPR-mutagenesis is mosaic, different cells can have different mutations, we used the Synthego ICE software tool^29^ to deconvolute the proportion and sequence of each indel mutation in each embryo (Supplemental Figure 1). Genotyping primers and gRNA sequences are in Table S3.

We only scored phenotypic data from embryos that had >40% mutation rate. For each gene, CRISPR-mutagenesis experiments were independently repeated at least twice in different batches of embryos, analyzing 5-15 individual mutant tadpoles per experiment. A candidate risk gene was determined to be likely-causative if more than 10% of mutant tadpoles had a tracheal or esophageal defect (LTEC, occluded esophagus, failed separation), compared to the baseline rate of <2% in control injected tadpoles.

## Results

A total of 185 individuals with EA/TEF were enrolled into the study, including 102 (55%) male and 83 (45%) female probands. Probands were between the ages of 2 days and 54.5 years with an average of 8.2 years old at enrollment (Table 1). The majority (52.4%) were type C EA/TEFs. Fifty-nine probands had isolated EA/TEF, and 126 probands had neurodevelopmental delay and/or at least one additional congenital anomaly and were classified as non-isolated. Of the non-isolated cases, the most common associated anomalies were congenital heart defects (65; 51.5%), skeletal defects (48; 38%) and renal defects (40; 31.7%). Other congenital anomalies included genitourinary defects (non-renal) (16; 12.7%), laryngotracheal defects (13; 10.3%), gastrointestinal defects (9; 7%), limb defects (7; 5.5%), neural tube defects (5; 3.9%), craniofacial defects (5; 3.9%) and other anomalies were seen in 12 probands (9.5%). Twenty-five probands (19.8%) had neurodevelopmental delay. The majority of probands were self-identified white (80.5%), and the remaining were Black/African American (8%), Asian (6%), more than one race (3.2%), or unknown (2.2%). One of the probands reported a family history of EA/TEF.

**Table 1.** Clinical table of 185 individuals enrolled into the study.

| Characteristics | N=185 |
| --- | --- |
| Mean age at enrollment (range) | 8.22 years (2 days-54.5 years) |
| *Sex* |  |
| Male | 102 (55%) |
| Female | 83 (45%) |
| *Race and ethnicity* |  |
| White | 149 (80.5%) |
| Black/African American | 15 (8%) |
| Asian | 11 (6%) |
| American Indian/Alaska Native | 0 |
| Native Hawaiian or Pacific Islander | 0 |
| More than one race | 6 (3.2%) |
| Unknown | 4 (2.2%) |
| Hispanic | 13 (7%) |
| Non-Hispanic | 169 (91.3%) |
| Unknown | 3 (1.6%) |
| *Type of EA/TEF* |  |
| Type A | 27 (14.5%) |
| Type B | 5 (2.7%) |
| Type C | 97 (52.4%) |
| Type D | 3 (1.6%) |
| Type H | 8 (4.3%) |
| Unknown | 45 (24.3%) |
| *Clinical presentation* |  |
| Isolated | 59 (32%) |
| Non-isolated | 126 (68%) |
| Cardiac defects | 65 (51.5%) |
| Skeletal defects | 48 (38%) |
| Renal defects | 40 (31.7%) |
| Neurodevelopmental delay | 21 (16.6%) |
| Genitourinary defects | 16 (12.7%) |
| Laryngotracheal defects | 13 (10.3%) |
| Gastrointestinal defects | 9 (7%) |
| Limb defects | 7 (5.5%) |
| Neural tube defects | 5 (3.9%) |
| Craniofacial defects | 5 (3.9%) |
| Other | 12 (9.5%) |

### Complex EA/TEF cases with additional anomalies have significant burden of *de novo* coding variants

We identified 249 *de novo* coding variants in 185 probands with EA/TEF (Table S4). The average number of *de novo* coding variants per proband is 1.35. We classified LGD and missense variants as protein altering variants. We identified 191 protein altering variants across all probands, including 47 in 59 isolated cases and 144 in 126 complex cases.

We performed a burden test for enrichment of *de novo* coding variants in all cases, isolated cases and complex cases respectively (Table 2a). The number of synonymous variants is close to expectation (fold=0.94, p=0.7). Overall, there is a significant burden of *de novo* protein altering variants (LGD or missense) (fold=1.22, p=4.2e-3). The burden is almost entirely observed in complex cases (fold=1.35, p=3.3e-4), as there is no evidence of *de novo* burden in isolated cases (fold=0.94, p=0.68). In complex cases (Table 2b), the burden of LGD variants is mostly in genes that are intolerant of loss of function variants (defined as gnomAD^16^ pLI≥0.5, “constrained genes”; fold=2.8, p=2.3e-3), similar to other developmental disorders^30^. The burden of *de novo* missense variants is also higher in constrained genes compared to non-constrained genes (fold=1.57 vs 1.22), although it is marginally significant in both constrained and non-constrained gene sets (p=3.3e-3 and 0.045, respectively). We estimate that about 38 genes carrying these variants in the complex cases are risk genes. Overall, *de novo* protein altering variants explain about 30% of population attributable risk of complex EA/TEF.

**Table 2a.**
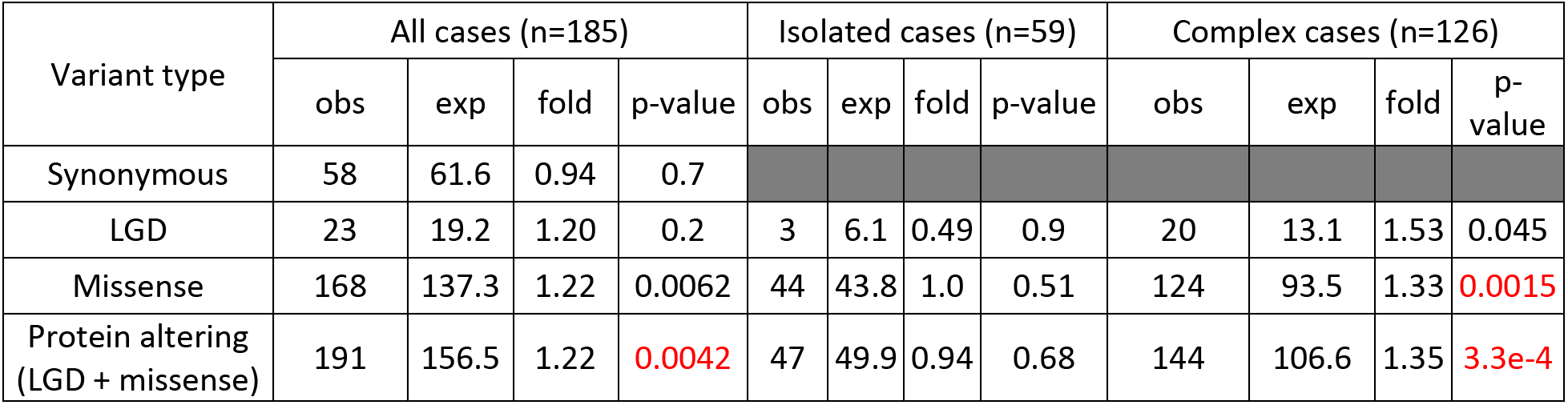
Burden of de novo variants in all cases. Burdens were calculated in all cases, isolated cases, and complex cases. Protein altering variants were defined as LGD and missense variants. LGD is likely gene disrupting. Obs is observed. Exp is expected. Table 2b. Burden of protein altering *de novo* variants in complex cases stratified by gene variant intolerance.

**Table 2b.**
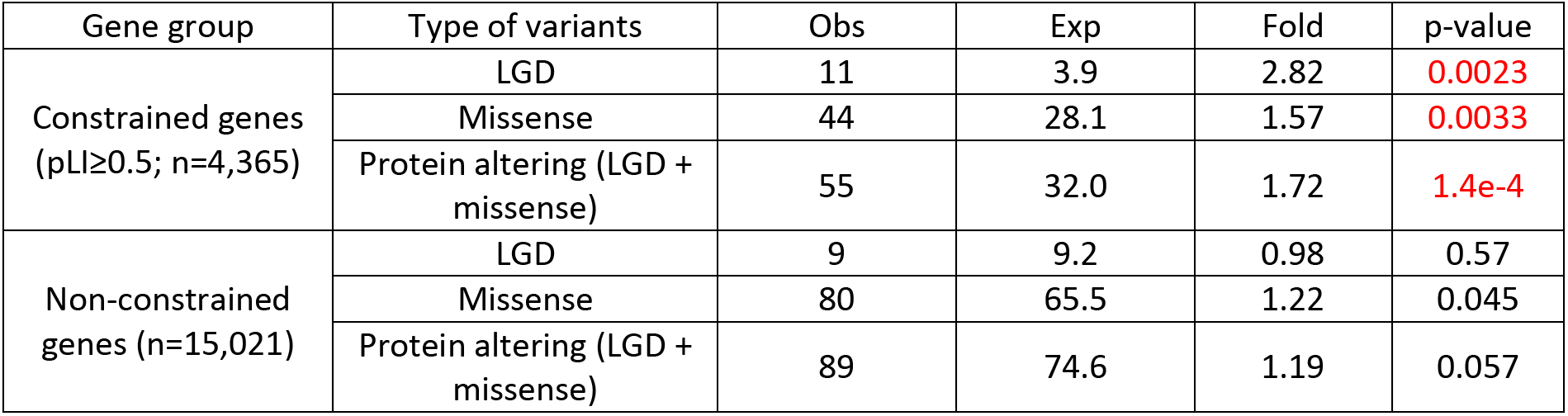
Burden of protein altering *de novo* variants in complex cases stratified by gene variant intolerance.

We assessed *de novo* protein coding variants for pathogenicity using the ACMG criteria^31^ (Table 3 and Table S5). Of the 185 cases, only two clearly had a molecular diagnosis consistent with the phenotype (*EFTU2* and *MYCN* associated with Mandibulofacial dysostosis, Guion-Almeida type (OMIM 610536) and Feingold syndrome (OMIM 164280), respectively). One individual with a *de novo* p.G365S *SMAD6* with a CADD score of 32 has a phenotype partially overlapping with conditions associated with *SMAD6* and may represent an expansion of the phenotypes associated with *SMAD6*. One individual with a de novo p.T647I variant in GLS with a CADD score of 27.1 has a phenotype that at the age of two does not overlap with OMIM 618339 with infantile cataracts, skin abnormalities, and intellectual disability. Of the 24 cases with *de novo* LGD variants, 21 were associated with complex phenotypes (Table 3).

**Table 3.**
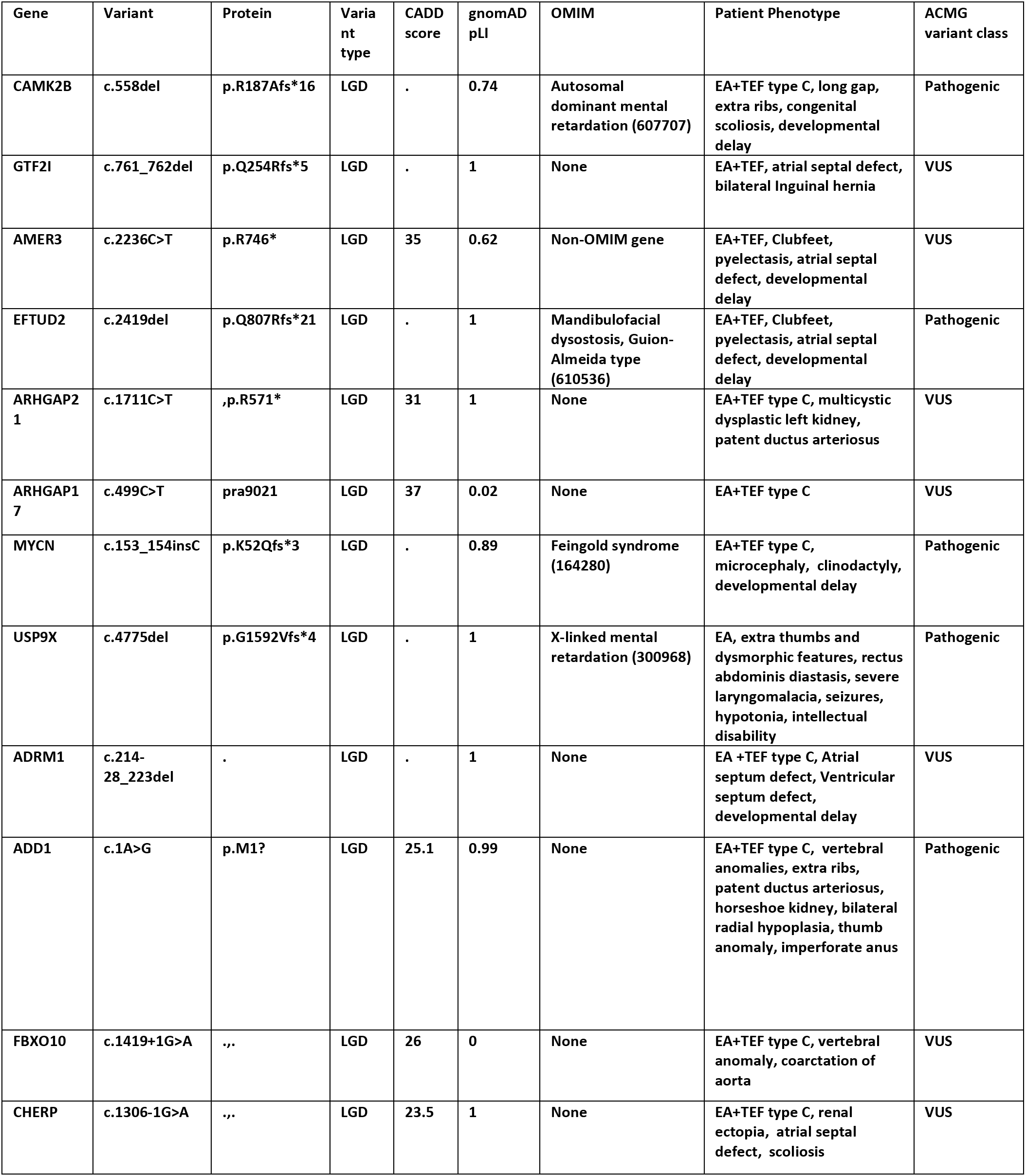
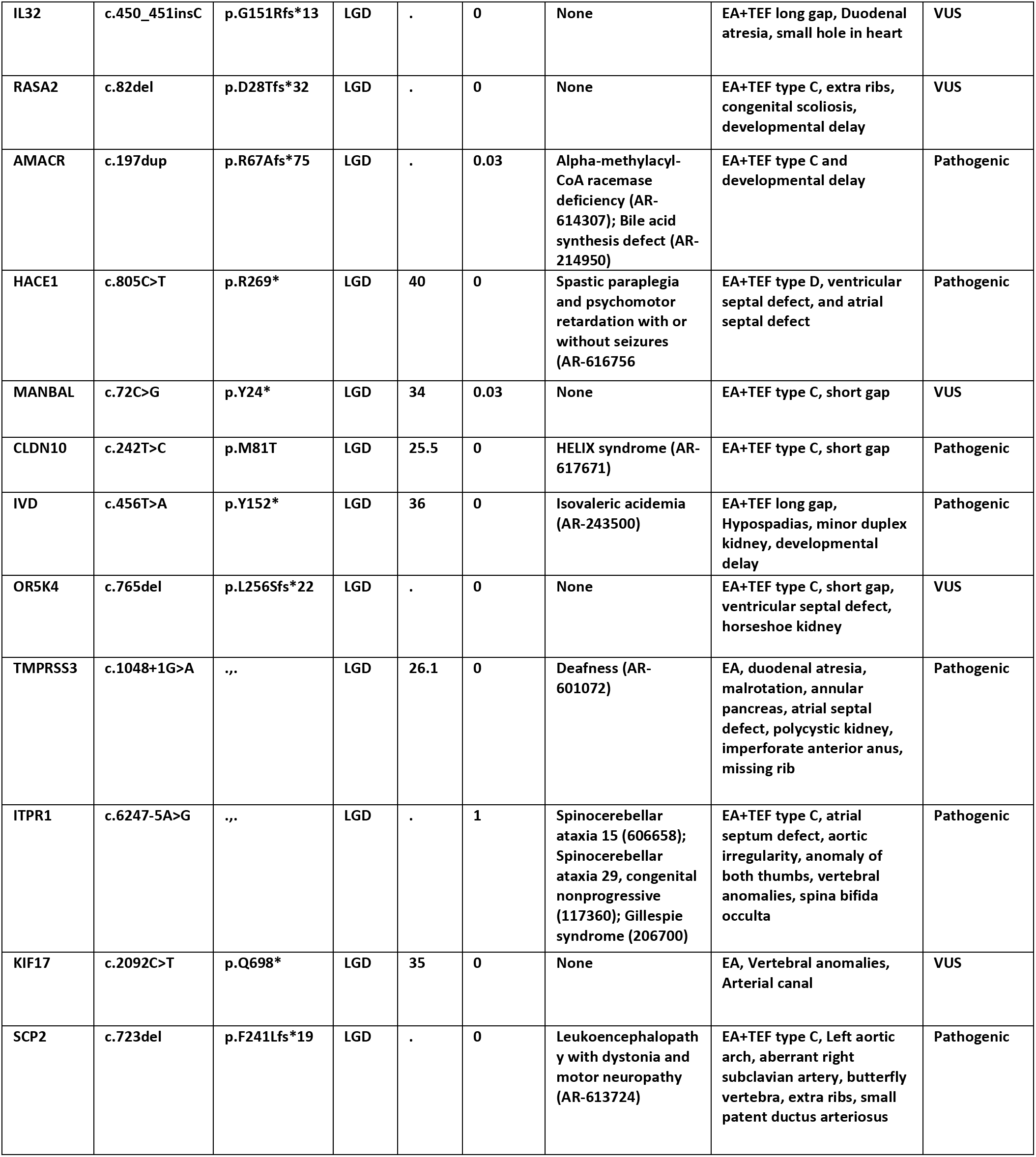
*De novo* LGD variants. LGD are likely gene disrupting. ACMG is American College of Medical Genetics. VUS is variant of uncertain significance.

### Protein altering variants in complex cases are involved in endosome trafficking and developmental pathways

While complex cases have a significant burden of *de novo* variants, no one gene harbors more than one LGD or missense *de novo* variant, making it impossible to identify individual risk genes with sufficient statistical support. To investigate the aggregate properties of risk genes, we performed pathway enrichment analysis on protein altering *de novo* variants in complex cases (n=126). We focused on GO (Gene Ontology) pathways and HPO (Human Phenotype Ontology) terms. To ensure sufficient statistical power, we only considered the pathways that are expected to have at least 2 protein altering variants by chance in 126 subjects. We compared the observed variants in each pathway to the expected number of variants estimated from background mutation rate, and tested the enrichment using a Poisson test. We corrected the multi-testing p-values to family wise error rate (FWER) based on simulations. Eight GO pathways and five HPO terms are enriched with protein altering *de novo* variants with FWER≤0.05 (Figure 1A, Table S6). The enriched GO pathways are related to autophagy processes, membrane regulation, and intracellular transport and localization, while the HPO terms are related to other developmental disorders (Figure 1B). A total of 86 genes are involved in at least one significant pathway. Fifty-five genes are involved in endocytosis and transcytosis pathways. Forty-five genes are involved in pathways related to other developmental disorders. The enrichment in GO pathways is mostly driven by *de novo* missense variants, whereas the enrichment in HPO terms is driven by both LGD and missense variants (Figure 1A). These findings are consistent with animal model studies in which pleiotropic signaling pathways and endosome-mediated epithelial remodeling are required for tracheoesophageal morphogenesis^2^.

**Figure 1.**
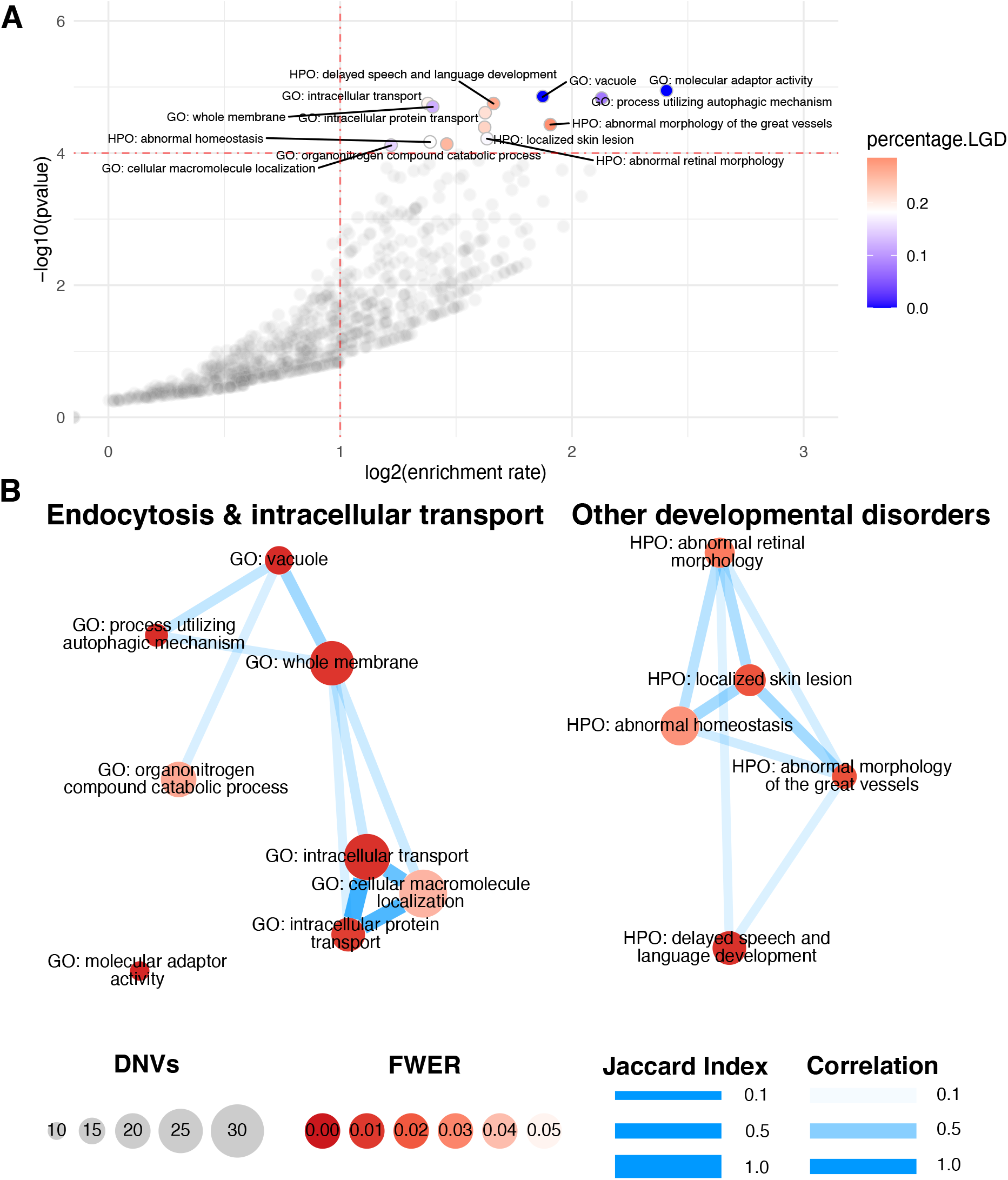
Pathway enrichment analysis. A) Volcano plot. Each dot represents a pathway. X-axis represents the enrichment rate in log scale, Y-axis is the Poisson test p-value in log10 scale. The horizontal dashed line marks family-wise error rate (FWER) of 0.05. Significant pathways (FWER<0.05) are colored by the percentage of LGD variants, other pathways are colored grey. B) Pathway overlaps. Each circle represents a pathway with FWER<0.05. Circle size is proportional to the number of observed *de novo* variants in the pathway; Circle color represents the FWER; Edge width is determined by the Jaccard Index between two pathways, and edge color represents the correlation coefficient of the two pathways under the null in simulations.

We also investigated the functional interactions among the genes (n=143) with protein altering *de novo* variants in complex cases. Based on stringDB (v11.0)^23^, the number of protein-protein interactions is significantly larger than expected (PPI enrichment p-value=0.0021, Fig.2).

**Figure 2.**
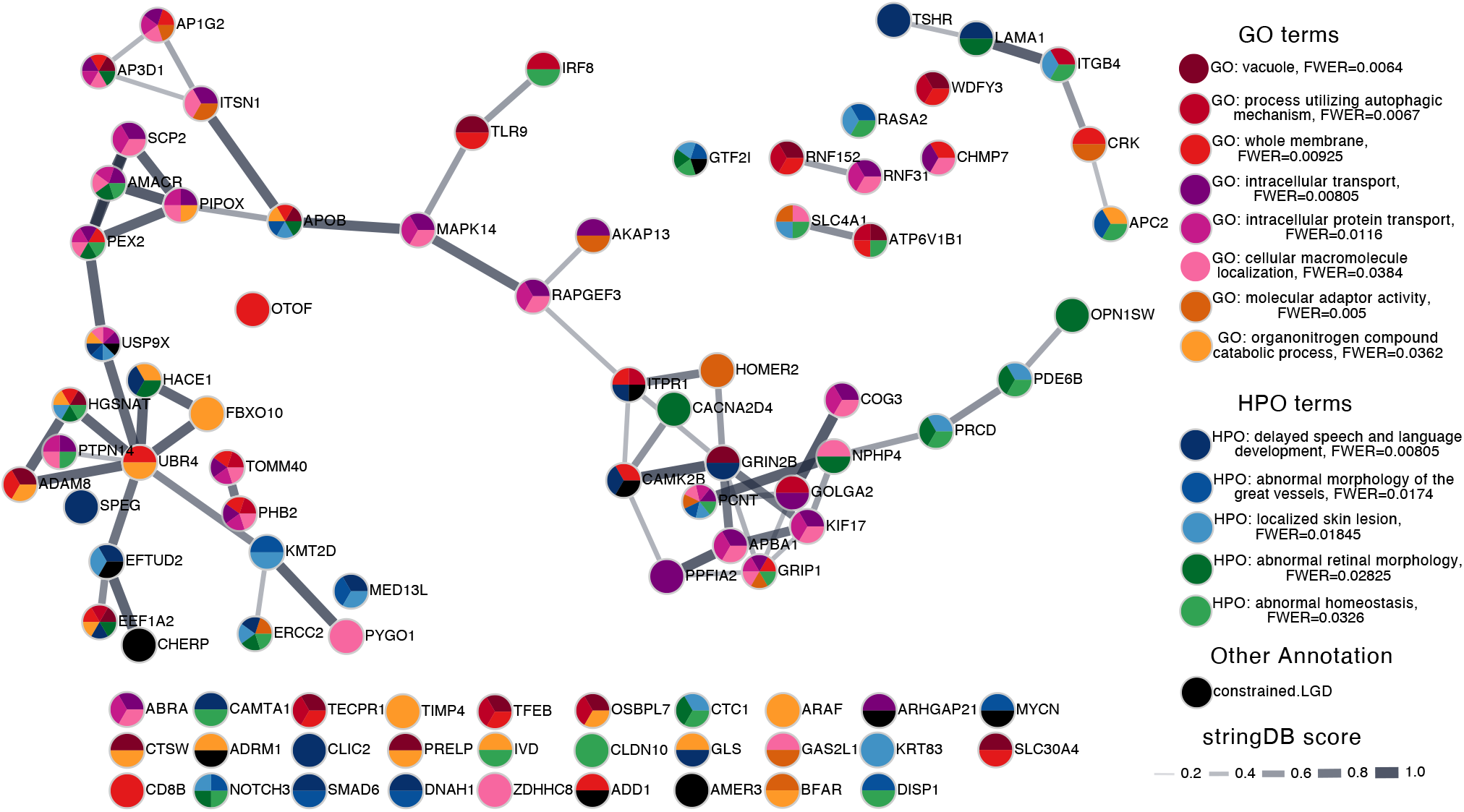
String-DB of LGD and missense genes in complex cases. Dots are colored to indicate whether it is involved in one of the significant pathways. Constrained genes (pLI ≥ 0.5) with LGD mutations were colored black. Edge width represents the stringDB score. Genes not involved in any of the annotation groups were not shown.

### CRISPR-mutation of candidate risk genes in *Xenopus* disrupts trachea-esophageal morphogenesis

The underlying biology of trachea and esophageal development is conserved between humans and other terrestrial vertebrates, and animal models have proven effective in assessing candidate risk variants from human patients^2^. We therefore turned to the rapid functional genomics possible in the amphibian *Xenopus,* which is increasingly being used to model human developmental disorders^32; 33^ including tracheoesophageal birth defects^12^.

We tested candidate risk variants by CRISPR-Cas9 mutagenesis of the orthologous genes in *Xenopus tropicalis,* assaying F0 mutant embryos rather than establishing multi-generational lines^34^ since this is faster and more closely mimics the *de novo* mutations in human EA/TEF patients. F0 mutagenesis results in embryos with a range of mosaic indel mutations. We found that F0 mutagenesis of *sox2*, a known EA/TEF risk gene in humans, resulted in a trachea-esophageal phenotype indistinguishable from F2 *sox2^-/-^* germline mutants with a failure of the foregut to separate into distinct esophagus and trachea (Figure 3B-C). Moreover, unlike human EA patients with heterozygous *SOX2^+/-^* mutations, heterozygous mouse and *Xenopus sox2^+/-^* mutants do not exhibit tracheoesophageal defects^8; 35^.

**Figure 3.**
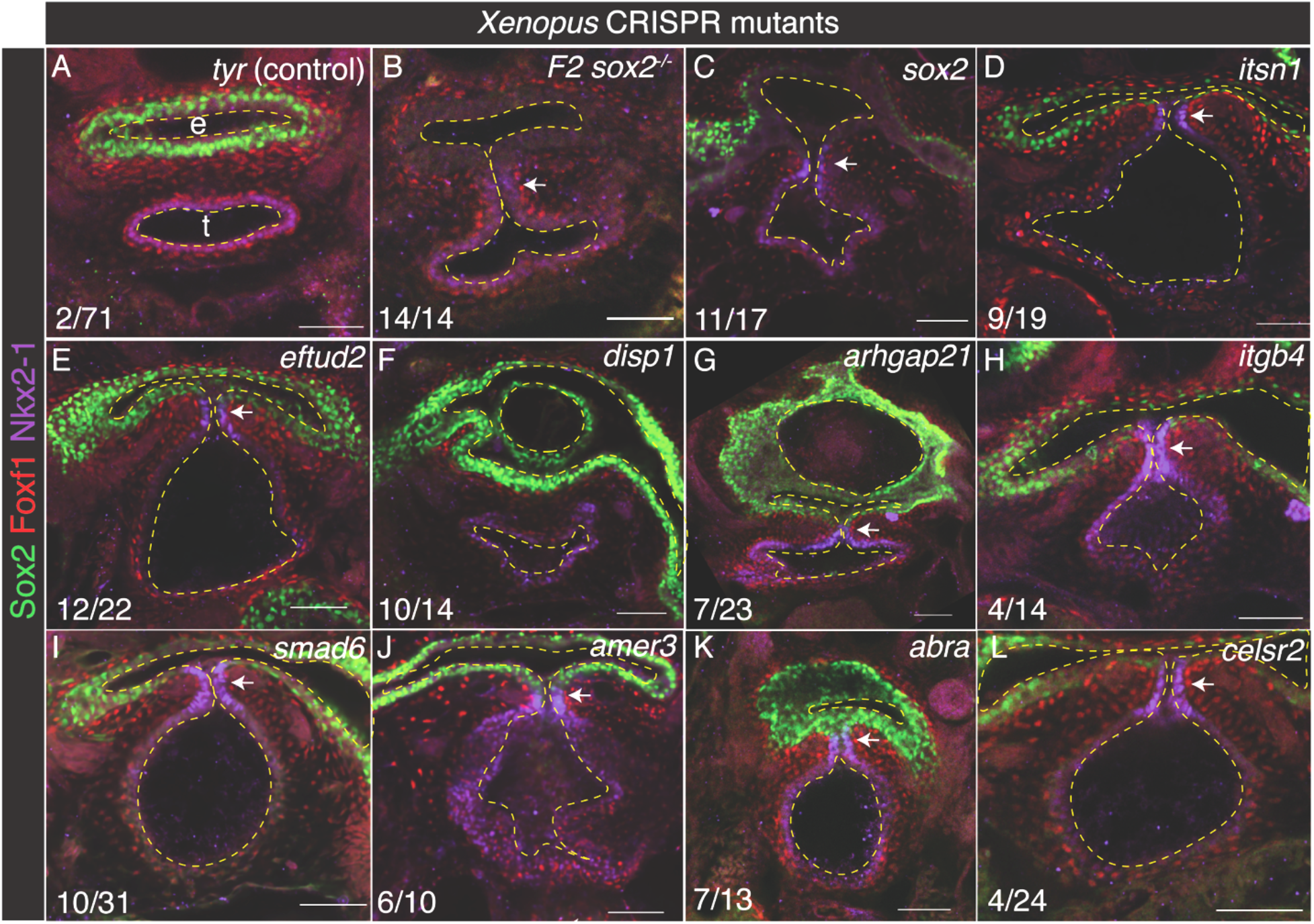
CRISPR-mutation of candidate risk genes in *Xenopus* disrupts trachea-esophagus morphogenesis. Representative confocal microscopy images of NF44 foregut from Xenopus CRISPR mutants. *Sox2* F0 CRISPR mutants (C) have the same trachea-esophageal phenotype as *sox2*^-/-^ F2 germline mutants (B), validating the F0 screen. Compared to control *tyr* mutations in which the trachea (t) and esophagus (e) have completely separated (A), mutation of 13/18 genes caused a failure of the foregut to separate into distinct trachea and esophagus (D-E,H-L) and/or resulted in a disrupted esophagus with multiple lumens (F,G). Dashed lines indicate the esophagus, trachea, and foregut lumens. Arrows point to a tracheoesophageal cleft. The number of embryos with a TED phenotype/total injected. Scale bars are 50 µm.

We prioritized and selected 18 candidate risk genes to test based on 1) the likelihood that the patient variant was damaging, 2) expression in the *Xenopus* and mouse fetal foregut and 3) the predicted function focusing on genes implicated in endosome trafficking or signaling pathways that pattern the fetal foregut (Table 4). Guide RNAs (gRNAs) were designed to generate loss of function (null) mutations or in a few cases where early embryonic lethality was predicted, a patient-like mutation targeting a conserved sequence near the patient’s variant. We genotyped each CRISPR-injected embryo and assessed the trachea-esophageal phenotype in embryos with >40% damaging indel mutations. At three days of development (stage NF44), when the trachea and esophagus have normally separated (Figure 3A), tadpoles were fixed and assessed by confocal immunostaining for Sox2+ esophageal epithelium, Nkx2-1+ tracheal epithelium and Foxf1+ foregut mesenchyme (Figure 3).

**Table 4.** EA/TEF candidate genes screened in Xenopus. TED=tracheoesophageal defect.

| Gene | Function | *Xenopus* TED frequency (n) | Co-occurring defects | % indels | Mutation type |
| --- | --- | --- | --- | --- | --- |
| *sox2* | Transcription factor | 100% (14) |  | 100% (germline) | Null |
| *sox2* | Transcription factor | 65% (17) | Microphthalmia | 91% | Null |
| *disp1* | Hedgehog signaling | 71% (14) |  | 62% | Null |
| *amer3* | Wnt signaling | 62% (21) |  | 57% | Null |
| *eftud2* | mRNA splicing | 55% (22) | Microphthalmia | 92% | Patient-like |
| *abra* | Rho signaling | 45% (20) | Craniofacial | 92% | Null |
| *itsn1* | Endocytosis | 47% (19) | Microcephaly | 71% | Null |
| *itsn1* | Endocytosis | 42% (43) |  | 72% | Patient-like |
| *apc2* | Wnt signaling | 37% (19) |  | 87% | Null |
| *smad6* | BMP signaling | 32% (31) | Craniofacial<br>Heart looping | 68% | Patient-like |
| *arhgap21* | Rho signaling | 30% (23) | Craniofacial | 86% | Null |
| *itgb4* | Integrin | 29% (14) | Heart looping | 76% | Null |
| *ap1g2* | Endocytosis | 24% (17) | Microphthalmia | 92% | Null |
| *rapgef3* | Ras signaling | 20% (5) |  | 91% | Null |
| *celsr2* | Wnt/PCP signaling | 17% (24) |  | 79% | Null |
| *ptpn14* | RTK signaling | 13% (23) |  | 94% | Null |
| *add1* | Cytoskeleton | 8% (24) | Craniofacial | 89% | Null |
| *map4k3* | MAPK signaling | 8% (13) |  | 71% | Null |
| *rab3gap2* | Endocytosis | 6% (18) |  | 81% | Null |
| *arhgap17* | Rho signaling | 0% (11) | Gut looping | 88% | Null |
| *pcdh1* | Cell-cell adhesion | 0% (7) |  | 47% | Null |
| *tyr* (control) | Pigmentation | 2% (71) |  | n/d | Null |

Thirteen of 18 genes screened exhibited defective trachea-esophageal development in >10% of mutated tadpoles (Table 4, Figure 3). The most common phenotype was a laryngo- tracheoesophageal cleft (LTEC) where the trachea and esophagus failed to separate near the larynx (e.g., *sox2*, *eftud2*, *itsn1*) (Figure 3C-E), or a disorganized esophageal epithelium, likely leading to esophageal atresia later in development (e.g., *arhgap21* and *disp1*) (Figure 3F-G). This failure to separate the embryonic foregut is a typical manifestation commonly observed in both mouse and *Xenopus* embryos with mutations in known EA risk genes^2^. Interestingly, several of the gene mutations also resulted in co-occurring defects in other organ systems like the EA/TEF human patients including microphthalmia, microcephaly, and craniofacial malformations. Notably, 5 genes are implicated in signaling pathways known to regulate foregut patterning (*amer3, apc2, celsr2, disp1, smad6*), while 5 other genes are implicated in endocytosis and/or intracellular trafficking (*abra, arhgap21, ap1g2, itsn1, rapgef3*) (Table 4).

## Discussion

In this study, we identified 249 *de novo* coding variants in 185 EA/TEF individuals, including 23 LGD variants and 168 missense variants. Only two cases were associated with pathogenic variants in genes previously established to cause EA/TEF, suggesting that most of our findings are identifying new genetic associations with EA/TEF. Protein altering *de novo* variants are enriched in complex cases. Consistent with previous studies of congenital anomalies, those variants showed greater enrichment in constrained genes. Pathway analysis showed that endocytosis, membrane regulation and intracellular trafficking related processes are enriched with protein altering variants. Considering recent findings in mouse and *Xenopus* that endosome-mediated epithelial remodeling acts downstream of Hedgehog-Gli signaling to regulate tracheoesophageal morphogenesis^12^, it is possible that disruption in endocytic vesicular trafficking may be a common mechanism in many EA/TEF patients.

Endocytic vesicular trafficking is regulated by small GTPases (Rab/Rho) that link endocytosis of membrane bound vesicles to the actin intracellular transport machinery, which moves vesicles to different subcellular compartments; to lysosomes in the case of autophagy, to different membrane domains in the case of recycling endosomes, from the Golgi and ER to the cell surface for maturation of membrane proteins, and from basal to apical membranes in the case of transcytosis^36–38^. Endocytic trafficking can influence morphogenesis in many ways; by changing cell shape, by dynamic remodeling of cell adhesion and junctional complexes, and by regulation of cell migration or cell signaling^39–43^. Moreover, one of the candidate genes that we tested, *ITSN1,* encodes a multidomain adaptor protein that coordinates the intracellular transport of endocytic vesicles^44^. *ITSN1* is also an autism risk loci and consistent with the neurodevelopmental disorders also present in the EA/TEF patient, Itsn1 is required for neural dendrite formation in rodents, where it physically interacts with core endocytic protein Dnm2 acting as a Cdc42-GEF to promote actin-mediated endosome transport^45; 46^. Thus, the finding that several candidate genes validated in Xenopus are implicated in endocytosis or GTPase activity (*abra, arhgap21, ap1g2, itsn1, rapgef3, rab3gap2),* suggests that the EA/TEF in the patients may have been due to disrupted foregut morphogenesis.

Our analysis also revealed that LGD and missense variants in complex cases are involved in other developmental disorders, suggesting disruptions to pleiotropic pathways with roles in multiple organ systems. Indeed, *Xenopus* mutagenesis validated several genes implicated in signaling pathways known to regulate foregut patterning as well as the development of other organ systems including *amer3, apc2,* and *celsr2* in the Wnt pathway, *smad6* and *sox2* in the BMP pathway, and *disp1* required for secretion of Hedgehog ligands. In the future, as more functional data are collected on EA/TEF risk variants, it may be possible to link distinct signaling pathways or cellar mechanisms such as endocytosis to different co-occurring anomalies in specific organ systems.

Overall, the genetics of EA/TEF is heterogenous. With 126 complex cases which are overall significantly enriched with *de novo* protein-altering variants, we did not find a gene with such variants in multiple cases. This indicates that the number of risk genes contributing through *de novo* variants is large. A sustained effort to expand the cohort with genome sequencing is critical to improve statistical power to identify new risk genes in human.

One interesting observation is that all CRISPR-generated Xenopus mutants had severe tracheoesophageal clefts rather than atresia or fistulas. We expect that this is because the CRISPR editing strategy results in high mutagenesis rates and often loss of function alleles resulting in more severe tracheoesophageal phenotypes, in contrast to the patients who have heterozygous variants. Indeed, in all most all reported cases where EA/TEF risk alleles have been modeled in mouse or *Xenopus*, heterozygous variants do not result in an EA/TEF phenotype, whereas null mutations exhibit a cleft with a single undivided foregut^2^. This difference could be due to hypomorphic human variants versus null alleles in animals. In humans, null alleles in pleiotropic developmental genes are likely to be embryonic lethal and may not be viable to term. An additional factor is likely to be the fact that animal models are inbred, whereas the humans have diverse genetic backgrounds, likely associated with modifying alleles. In the future it will be important to test these possibilities with the exact patient alleles in animal models to obtain a better assess the genotype-phenotype relationship of these conditions.

## Limitations

Our study had limited statistical power to identify individual risk genes of EA/TEF based solely upon the human genetic studies due to the limited sample size. Collaborative human genetic studies of EA/TEF will be necessary to increase those sample sizes and better understand the spectrum of phenotypes associated with each gene.

However, even with a modest human sample size with only a single human with a *de novo* variant in the gene, we demonstrate the ability to effectively select disease causing variants and functionally confirm the majority of the candidate genes using a moderate throughput F0 mutagenesis system. The combination of human genetics and model organism modeling is powerful for rare human genetic conditions associated with morphological defects. By examining pathways common across genes, we implicate endocytosis, membrane regulation and intracellular trafficking in tracheoesophageal development, and these same processes are likely related to other congenital anomalies and neurodevelopmental disorders.

## Data and Code availability

Code for pathway enrichment analysis of *de novo* variants with family wise error rate estimation is available on GitHub: https://github.com/ShenLab/pathways

Whole genome sequencing data of the cases is available on dbGaP (phs002161).

## Data Availability

Code for pathway enrichment analysis of de novo variants with family wise error rate estimation is available on GitHub: https://github.com/ShenLab/pathways
Whole genome sequencing data of the cases is available on dbGaP (phs002161).

## Acknowledgements

We would like to thank the patients and their families who participated in the study and the TOFS UK organization for their support of the study. We thank Patricia Lanzano, Jiangyuan Hu, Liyong Deng, and Charles LeDuc from Columbia University for technical assistance. We would like to thank the study coordinators Gentry Wools (UT-Southwest), Amanda Schreibeis (Cincinnati Children’s Hospital) and Andrew Mason (Oregon Health and Science University) for their assistance. We also thank Dr. Na Zhu, Dr. Xueya Zhou, and other members of Chung and Shen labs for helpful discussions. The whole genome sequencing data were generated through NIH Gabriella Miller Kids First Pediatric Research Program (X01HL145692 and X01HD100705). Microscopy was performed at the CCHMC Confocal Imaging Core. The work was support by NIH grants P01HD093363 (AMZ, YS, and WKC), R01GM120609 (GZ, YS), and R03HL147197 (YS). NAE is supported by a Canadian Institutes of Health Research postdoctoral fellowship. We acknowledge Marko Horb and Danielle Jordan at the National *Xenopus* Resource for generating *sox2* germline mutant *Xenopus* tadpoles for this study (RRID:SCR_013731).

## Author contributions

WKC, YS and AMZ had full access to all of the data in the study and take responsibility for the integrity of the data and the accuracy of the data analysis. Concept and design: WKC, YS and AMZ. Acquisition, analysis, or interpretation of data: GZ, PA, NAE, JJH, CF, PK, WM, JK, MEF, DS, EF, AK, SF, APK, AMZ, YS and WKC. Drafting of the manuscript: GZ, PA, NAE, AMZ, YS and WKC. Critical revision of the manuscript for important intellectual content: GZ, PA, NAE, JJH, CF, PK, WM, JK, MEF, DS, EF, AK, SF, APK, AMZ, YS and WKC. Statistical analysis: GZ, PA, NAE, AMZ, and YS. *Xenopus* experimental design: NAE, AK. *Xenopus* experiment: NAE, AK, SF, APK. Supervision: AMZ, YS and WKC. The authors read and approved the final manuscript.

## References

1. Nassar, N., Leoncini, E., Amar, E., Arteaga-Vazquez, J., Bakker, M.K., Bower, C., Canfield, M.A., Castilla, E.E., Cocchi, G., Correa, A., et al. (2012). Prevalence of esophageal atresia among 18 international birth defects surveillance programs. Birth Defects Res A 94, 893–899.

2. Edwards, N.A., Shacham-Silverberg, V., Weitz, L., Kingma, P.S., Shen, Y., Wells, J.M., Chung, W.K., and Zorn, A.M. (2021). Developmental basis of trachea-esophageal birth defects. Dev Biol 477, 85–97.

3. van Lennep, M., Singendonk, M.M.J., Dall’Oglio, L., Gottrand, F., Krishnan, U., Terheggen-Lagro, S.W.J., Omari, T.I., Benninga, M.A., and van Wijk, M.P. (2019). Oesophageal atresia. Nat Rev Dis Primers 5, 26.

4. Stoll, C., Alembik, Y., Dott, B., and Roth, M.P. (2017). Associated anomalies in cases with esophageal atresia. Am J Med Genet A 173, 2139–2157.

5. Shaw-Smith, C. (2010). Genetic factors in esophageal atresia, tracheo-esophageal fistula and the VACTERL association: roles for FOXF1 and the 16q24.1 FOX transcription factor gene cluster, and review of the literature. Eur J Med Genet 53, 6–13.

6. Li, Y., Litingtung, Y., Ten Dijke, P., and Chiang, C. (2007). Aberrant Bmp signaling and notochord delamination in the pathogenesis of esophageal atresia. Dev Dyn 236, 746–754.

7. Que, J., Choi, M., Ziel, J.W., Klingensmith, J., and Hogan, B.L. (2006). Morphogenesis of the trachea and esophagus: current players and new roles for noggin and Bmps. Differentiation 74, 422–437.

8. Que, J., Okubo, T., Goldenring, J.R., Nam, K.T., Kurotani, R., Morrisey, E.E., Taranova, O., Pevny, L.H., and Hogan, B.L. (2007). Multiple dose-dependent roles for Sox2 in the patterning and differentiation of anterior foregut endoderm. Development 134, 2521–2531.

9. Rankin, S.A., Han, L., McCracken, K.W., Kenny, A.P., Anglin, C.T., Grigg, E.A., Crawford, C.M., Wells, J.M., Shannon, J.M., and Zorn, A.M. (2016). A Retinoic Acid-Hedgehog Cascade Coordinates Mesoderm-Inducing Signals and Endoderm Competence during Lung Specification. Cell Rep 16, 66–78.

10. Gordon, C.T., Petit, F., Oufadem, M., Decaestecker, C., Jourdain, A.S., Andrieux, J., Malan, V., Alessandri, J.L., Baujat, G., Baumann, C., et al. (2012). EFTUD2 haploinsufficiency leads to syndromic oesophageal atresia. J Med Genet 49, 737–746.

11. Wang, J., Ahimaz, P.R., Hashemifar, S., Khlevner, J., Picoraro, J.A., Middlesworth, W., Elfiky, M.M., Que, J., Shen, Y., and Chung, W.K. (2021). Novel candidate genes in esophageal atresia/tracheoesophageal fistula identified by exome sequencing. Eur J Hum Genet 29, 122–130.

12. Nasr, T., Mancini, P., Rankin, S.A., Edwards, N.A., Agricola, Z.N., Kenny, A.P., Kinney, J.L., Daniels, K., Vardanyan, J., Han, L., et al. (2019). Endosome-Mediated Epithelial Remodeling Downstream of Hedgehog-Gli Is Required for Tracheoesophageal Separation. Dev Cell 51, 665–674 e666.

13. Qi, H., Yu, L., Zhou, X., Wynn, J., Zhao, H., Guo, Y., Zhu, N., Kitaygorodsky, A., Hernan, R., Aspelund, G., et al. (2018). De novo variants in congenital diaphragmatic hernia identify MYRF as a new syndrome and reveal genetic overlaps with other developmental disorders. PLoS Genet 14, e1007822.

14. Richter, F., Morton, S.U., Kim, S.W., Kitaygorodsky, A., Wasson, L.K., Chen, K.M., Zhou, J., Qi, H., Patel, N., DePalma, S.R., et al. (2020). Genomic analyses implicate noncoding de novo variants in congenital heart disease. Nat Genet 52, 769–777.

15. Poplin, R., Chang, P.C., Alexander, D., Schwartz, S., Colthurst, T., Ku, A., Newburger, D., Dijamco, J., Nguyen, N., Afshar, P.T., et al. (2018). A universal SNP and small-indel variant caller using deep neural networks. Nat Biotechnol 36, 983–987.

16. Karczewski, K.J., Francioli, L.C., Tiao, G., Cummings, B.B., Alfoldi, J., Wang, Q., Collins, R.L., Laricchia, K.M., Ganna, A., Birnbaum, D.P., et al. (2020). The mutational constraint spectrum quantified from variation in 141,456 humans. Nature 581, 434–443.

17. Lek, M., Karczewski, K.J., Minikel, E.V., Samocha, K.E., Banks, E., Fennell, T., O’Donnell-Luria, A.H., Ware, J.S., Hill, A.J., Cummings, B.B., et al. (2016). Analysis of protein-coding genetic variation in 60,706 humans. Nature 536, 285–291.

18. Jaganathan, K., Kyriazopoulou Panagiotopoulou, S., McRae, J.F., Darbandi, S.F., Knowles, D., Li, Y.I., Kosmicki, J.A., Arbelaez, J., Cui, W., Schwartz, G.B., et al. (2019). Predicting Splicing from Primary Sequence with Deep Learning. Cell 176, 535–548 e524.

19. Samocha, K.E., Robinson, E.B., Sanders, S.J., Stevens, C., Sabo, A., McGrath, L.M., Kosmicki, J.A., Rehnstrom, K., Mallick, S., Kirby, A., et al. (2014). A framework for the interpretation of de novo mutation in human disease. Nat Genet 46, 944–950.

20. Ware, J.S., Samocha, K.E., Homsy, J., and Daly, M.J. (2015). Interpreting de novo Variation in Human Disease Using denovolyzeR. Curr Protoc Hum Genet 87, 7 25 21–27 25 15.

21. Mootha, V.K., Lindgren, C.M., Eriksson, K.F., Subramanian, A., Sihag, S., Lehar, J., Puigserver, P., Carlsson, E., Ridderstrale, M., Laurila, E., et al. (2003). PGC-1alpha-responsive genes involved in oxidative phosphorylation are coordinately downregulated in human diabetes. Nat Genet 34, 267–273.

22. Subramanian, A., Tamayo, P., Mootha, V.K., Mukherjee, S., Ebert, B.L., Gillette, M.A., Paulovich, A., Pomeroy, S.L., Golub, T.R., Lander, E.S., et al. (2005). Gene set enrichment analysis: a knowledge-based approach for interpreting genome-wide expression profiles. Proc Natl Acad Sci U S A 102, 15545–15550.

23. Szklarczyk, D., Gable, A.L., Lyon, D., Junge, A., Wyder, S., Huerta-Cepas, J., Simonovic, M., Doncheva, N.T., Morris, J.H., Bork, P., et al. (2019). STRING v11: protein-protein association networks with increased coverage, supporting functional discovery in genome-wide experimental datasets. Nucleic Acids Res 47, D607–D613.

24. Shannon, P., Markiel, A., Ozier, O., Baliga, N.S., Wang, J.T., Ramage, D., Amin, N., Schwikowski, B., and Ideker, T. (2003). Cytoscape: a software environment for integrated models of biomolecular interaction networks. Genome Res 13, 2498–2504.

25. Lane, M., and Khokha, M.K. (2021). Obtaining Xenopus tropicalis Embryos by In Vitro Fertilization. Cold Spring Harb Protoc.

26. Lane, M., and Khokha, M.K. (2021). Obtaining Xenopus tropicalis Embryos by Natural Mating. Cold Spring Harb Protoc.

27. Moreno-Mateos, M.A., Vejnar, C.E., Beaudoin, J.D., Fernandez, J.P., Mis, E.K., Khokha, M.K., and Giraldez, A.J. (2015). CRISPRscan: designing highly efficient sgRNAs for CRISPR-Cas9 targeting in vivo. Nat Methods 12, 982–988.

28. Fortriede, J.D., Pells, T.J., Chu, S., Chaturvedi, P., Wang, D., Fisher, M.E., James-Zorn, C., Wang, Y., Nenni, M.J., Burns, K.A., et al. (2020). Xenbase: deep integration of GEO & SRA RNA-seq and ChIP-seq data in a model organism database. Nucleic Acids Res 48, D776–D782.

29. Hsiau, T., Conant, D., Rossi, N., Maures, T., Waite, K., Yang, J., Joshi, S., Kelso, R., Holden, K., Enzmann, B.L., et al. (2019). Inference of CRISPR Edits from Sanger Trace Data. bioRxiv, 251082.

30. Satterstrom, F.K., Kosmicki, J.A., Wang, J., Breen, M.S., De Rubeis, S., An, J.Y., Peng, M., Collins, R., Grove, J., Klei, L., et al. (2020). Large-Scale Exome Sequencing Study Implicates Both Developmental and Functional Changes in the Neurobiology of Autism. Cell 180, 568–584 e523.

31. Richards, S., Aziz, N., Bale, S., Bick, D., Das, S., Gastier-Foster, J., Grody, W.W., Hegde, M., Lyon, E., Spector, E., et al. (2015). Standards and guidelines for the interpretation of sequence variants: a joint consensus recommendation of the American College of Medical Genetics and Genomics and the Association for Molecular Pathology. Genet Med 17, 405–424.

32. Fakhro, K.A., Choi, M., Ware, S.M., Belmont, J.W., Towbin, J.A., Lifton, R.P., Khokha, M.K., and Brueckner, M. (2011). Rare copy number variations in congenital heart disease patients identify unique genes in left-right patterning. Proc Natl Acad Sci U S A 108, 2915–2920.

33. Willsey, H.R., Exner, C.R.T., Xu, Y., Everitt, A., Sun, N., Wang, B., Dea, J., Schmunk, G., Zaltsman, Y., Teerikorpi, N., et al. (2021). Parallel in vivo analysis of large-effect autism genes implicates cortical neurogenesis and estrogen in risk and resilience. Neuron 109, 788–804 e788.

34. Bhattacharya, D., Marfo, C.A., Li, D., Lane, M., and Khokha, M.K. (2015). CRISPR/Cas9: An inexpensive, efficient loss of function tool to screen human disease genes in Xenopus. Dev Biol 408, 196–204.

35. Trisno, S.L., Philo, K.E.D., McCracken, K.W., Cata, E.M., Ruiz-Torres, S., Rankin, S.A., Han, L., Nasr, T., Chaturvedi, P., Rothenberg, M.E., et al. (2018). Esophageal Organoids from Human Pluripotent Stem Cells Delineate Sox2 Functions during Esophageal Specification. Cell Stem Cell 23, 501–515 e507.

36. Grant, B.D., and Donaldson, J.G. (2009). Pathways and mechanisms of endocytic recycling. Nat Rev Mol Cell Biol 10, 597–608.

37. Naslavsky, N., and Caplan, S. (2018). The enigmatic endosome - sorting the ins and outs of endocytic trafficking. J Cell Sci 131.

38. Serra, N.D., and Sundaram, M.V. (2021). Transcytosis in the development and morphogenesis of epithelial tissues. EMBO J 40, e106163.

39. Lee, J.Y., and Harland, R.M. (2010). Endocytosis is required for efficient apical constriction during Xenopus gastrulation. Curr Biol 20, 253–258.

40. Bruser, L., and Bogdan, S. (2017). Adherens Junctions on the Move-Membrane Trafficking of E-Cadherin. Cold Spring Harb Perspect Biol 9.

41. Mathew, R., Rios-Barrera, L.D., Machado, P., Schwab, Y., and Leptin, M. (2020). Transcytosis via the late endocytic pathway as a cell morphogenetic mechanism. EMBO J 39, e105332.

42. Kowalczyk, I., Lee, C., Schuster, E., Hoeren, J., Trivigno, V., Riedel, L., Gorne, J., Wallingford, J.B., Hammes, A., and Feistel, K. (2021). Neural tube closure requires the endocytic receptor Lrp2 and its functional interaction with intracellular scaffolds. Development 148.

43. Yoon, J., Garo, J., Lee, M., Sun, J., Hwang, Y.S., and Daar, I.O. (2021). Rab11fip5 regulates telencephalon development via ephrinB1 recycling. Development 148.

44. Hussain, N.K., Yamabhai, M., Ramjaun, A.R., Guy, A.M., Baranes, D., O’Bryan, J.P., Der, C.J., Kay, B.K., and McPherson, P.S. (1999). Splice variants of intersectin are components of the endocytic machinery in neurons and nonneuronal cells. J Biol Chem 274, 15671–15677.

45. Yu, Y., Chu, P.Y., Bowser, D.N., Keating, D.J., Dubach, D., Harper, I., Tkalcevic, J., Finkelstein, D.I., and Pritchard, M.A. (2008). Mice deficient for the chromosome 21 ortholog Itsn1 exhibit vesicle-trafficking abnormalities. Hum Mol Genet 17, 3281–3290.

46. Gryaznova, T., Gubar, O., Burdyniuk, M., Kropyvko, S., and Rynditch, A. (2018). WIP/ITSN1 complex is involved in cellular vesicle trafficking and formation of filopodia-like protrusions. Gene 674, 49–56.

