## Supplementary Figures for "Identification and validation of novel candidate risk genes in endocytic vesicular trafficking associated with esophageal atresia and tracheoesophageal fistulas"

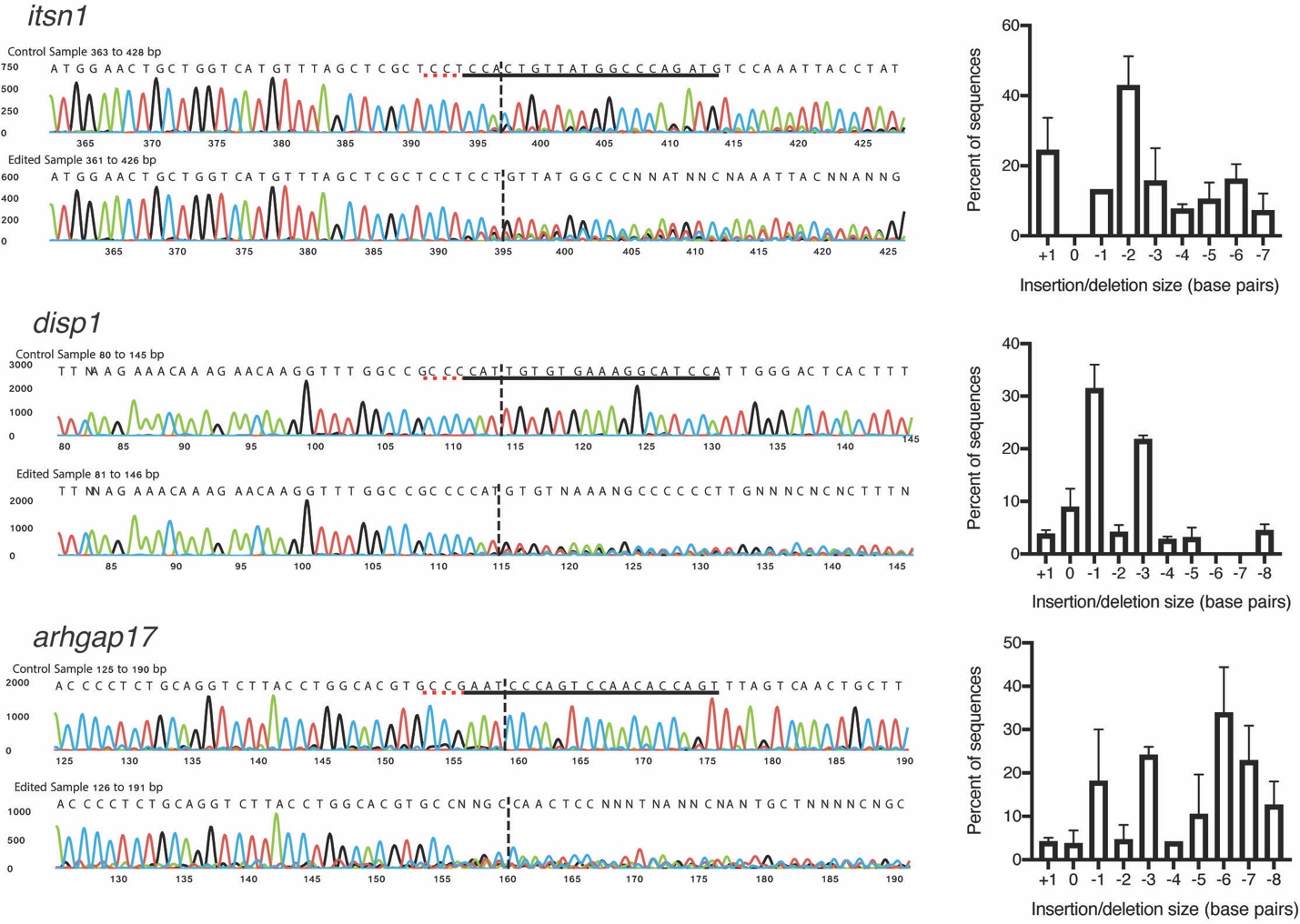

**Supplemental Figure 1**. Deconvolution of CRISPR-mediated indels in *Xenopus* embryos. CRISPR-Cas9-injected *Xenopus* embryos were genotyped by PCR amplification of the target region followed by Sanger sequencing. Sanger sequencing traces were analyzed using ICE software (Synthego, USA) to deconvolute the proportion and sequence of each indel mutation in each embryo. Representative sample traces are shown for gene editing of *itsn1*, *disp1*, and *arhgap17* (traces generated by ICE). The gRNA sequence is indicated by black lines and the PAM sequence by dotted red lines. Dashed vertical black lines indicate the predicted location of the Cas9 cut site. Bar graphs indicate the percent of sequences with the indicated insertion or deletion size (in base pairs) ± standard error of the mean (n=5-10 embryos analyzed per graph). F0 CRISPR editing of embryos results in mosaic indels however each gRNA tends to generate similar mutation profiles in embryos within a given experiment (e.g. *itsn1* gRNA tends to produce -2 and +1 bp indels).
